# Performance of Valved Respirators to Reduce Respiratory Particles Generated by Speaking

**DOI:** 10.1101/2021.10.23.21265427

**Authors:** Jessica M. Hazard, Christopher D. Cappa

## Abstract

Wearing of face coverings serves two purposes: reducing the concentration of ambient particles inhaled and reducing the emission of respiratory particles generated by the wearer. The efficiency of different face coverings depends on the material, design, and fit. Face coverings such as N95 respirators, when worn properly, are highly efficient at filtering ambient particles during inhalation. Some N95 respirators, as well as other face covering types, include a one-way valve to allow easier exhalation while still maintaining high filtration efficiency towards ambient particles. The extent to which these valves decrease filtration of emitted respiratory particles is, however, not well established. Here, we show that different valved N95s exhibit highly variable filtration efficiencies for exhaled respiratory particles. As such, valved N95s do not provide reliable source control of respired particles and their use should be discouraged in situations where such source control is needed.

## MAIN TEXT

The effectiveness of face coverings for reducing emission of inhaled ambient particles and exhaled respiratory particles varies with the type of face coverings. In many settings, the primary purpose of face coverings, generically referred to as masks, is to protect the wearer from inhalation of exogenous ambient particles that might be toxic or otherwise unhealthy.^1^ Generally, cloth masks and medical procedure masks do not provide the same level of reduction as a well-fit respirator (e.g., N95 facepieces) for inhaled particles.^2-5^ Some N95 respirators, as well as other types of face coverings (referred to here generically as masks), include an exhalation valve, the purpose of which is to facilitate easier breathing and reduce humidity and temperature inside the mask deadspace while still providing protection to the wearer against ambient particles.^6, 7^ The inclusion of an exhalation valve makes sense if the primary purpose is to protect the wearer, so long as it does not affect the mask filtration efficiency towards ambient particles. However, as the ongoing COVID-19 pandemic has made clear, masks also provide an important other function, namely source control via reduction of the emission of potentially infectious respiratory particles^8, 9^ that are produced during breathing, speaking, coughing, or sneezing.^10, 11^

In this context, it is critical to understand the extent to which an exhalation valve reduces mask efficiency towards exhaled respired particles and to compare their performance with other mask types. Staymates (2021) provides qualitative evidence that valved N95 respirators lead to excessive escape of respired particles and therefore a substantial reduction in the filtration efficiency.^12^ National Institute for Occupational Safety and Health (NIOSH) researchers performed experiments using test aerosol to challenge various valved respirators firmly sealed around their edges to a surface. In contrast to Staymates,^12^ NIOSH found relatively high efficiencies, despite the presence of the valve, although it is possible that the valves in the respirators used may have remained mostly closed during testing leading to artificially high efficiencies.^13^ Additionally, these measurements considered performance under ideal conditions (perfect sealing) and did not address performance when worn by people. Asadi et al. (2020) provided measurements of the effectiveness of a vented N95 respirator towards respiratory particles when worn by people, finding reasonably good performance.^14^ However, their measurements were limited to two people only and one respirator type. Also, they only measured particle emissions in the forward direction and may have undersampled any particles that escaped through the valve.

To provide the public with clear guidance regarding appropriate mask wearing to reduce both inhaled and exhaled particle concentrations requires clear understanding of the reduction afforded by valved respirators when worn by actual people while speaking. Speaking is one of the most common respiratory particle generating processes that leads to emission of particles at about 10x the rate of breathing.^10^ Here, we address this issue by making measurements of the reduction in respiratory particle concentrations generated by people when speaking afforded by wearing of different masks, including readily available (in the U.S.) valved N95 respirators.

## MATERIALS AND METHODS

Following from the methods used by Cappa et al. (2021) and associated other works,^10, 14-16^ we measured the concentrations of respiratory particles emitted while speaking by 10 individuals ranging in age from 20-43 with four self-identified females and six self-identified males. The University of California Davis Institutional Review Board approved this study (IRB# 844,369–4), and all research was performed in accordance with relevant guidelines and regulations of the Institutional Review Board. The participants spoke the *Rainbow Passage* while either not wearing or wearing one of four face coverings: a surgical procedure mask (ValuMax, Model: 5130E-SB), a valved 3M N95 respirator (Model: 8511), the same 3M 8511 N95 but with the valve taped over in the mask interior, or a valved Milwaukee N95 respirator (Model: 48-73-4011). These particular valved respirators were selected as they are readily available to the public in the U.S. Participants were provided instructions for and guided towards proper wearing of the masks but no formal fit test was conducted; the intent here is to consider masks as they might be worn by the public. To reduce the potential for sticking of the N95 respirator valves the valve flaps were gently pushed out prior to the initial wearing.

Participants spoke with their face and the sides of the face coverings inside the outer circumference of a large (30 cm diameter) funnel from which an Aerodynamic Particle Sizer (APS; TSI, Inc, 5 lpm) and a Condensation Particle Counter (CPC; TSI, Inc., 0.3 lpm) continuously sampled along with an excess flow of 19.7 lpm, such that the total flow into the funnel was 25 lpm. The APS characterizes size distributions and concentrations of particles having diameters >0.5 microns while the CPC measures the concentration of all particles >0.01 microns. The stopping distance of 1 micron particles that escape from the mask sides is << 1 cm and thus these will be entrained into the airflow passing into the funnel. A laminar flow hood (Air Science, PURAIR FLOW-48) housed the sampling funnel and provided HEPA filtered air such that background particle concentrations were negligible. Figure 1 shows the experimental setup.

**Figure 1:**
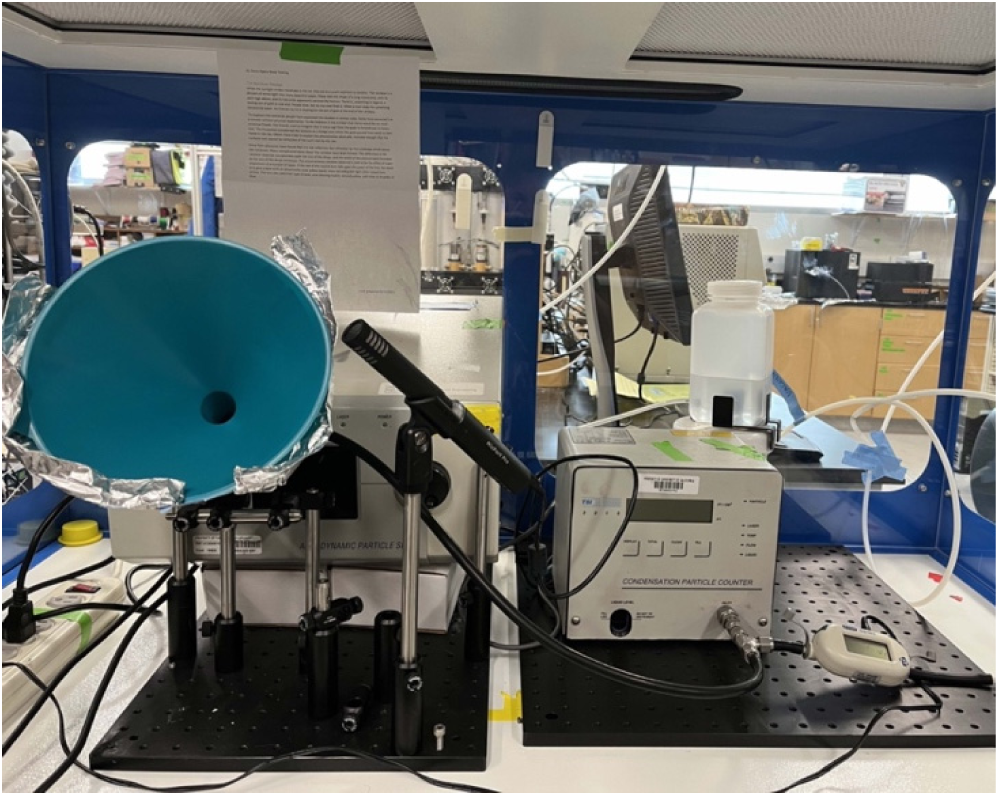
Experimental setup to measure the concentration of respiratory particles emitted while speaking.

Each participant performed two non-sequential replicates for each condition using the same mask and the order of tests was varied semi-randomly between participants. One participant repeated these tests using different masks (e.g., multiple readings wearing different 3M 8511 respirators) to help establish whether between-participant differences derive primarily from differences in how the individuals wore the masks and spoke versus from differences in the individual masks. This participant also performed the tasks wearing a non-valved N95 respirator (3M, Model Aura 9205+). The ratio (*R*_mask_) between the particle concentration measured with wearing of a given mask and without provides a measure of the mask efficiency (*η*_*mask*_ = 1 − *R*_*mask*_) for reducing emission of respiratory particles.

## RESULTS AND DISCUSSION

With no face covering, measured particle concentrations and size distributions were consistent with previous observations,^10, 11, 15-17^ with the CPC measuring on average 33x as many particles as the APS, indicating that particles <0.5 microns dominate the overall number. Comparing the observations across all participants, the median (or geometric mean) mask-to-no mask ratio for all particles varied substantially between face covering types, at 0.48 (0.40) for the Milwaukee, 0.12 (0.10) for the 3M 8511, 0.07 (0.07) for the taped 3M 8511, and 0.13 (0.10) for the surgical masks (*Figure 2*a). The results for particles >0.5 microns were similar (not shown). The multiple repeats by the one participant wearing different individual masks yielded similar results (*Figure 2*b).

**Figure 2:**
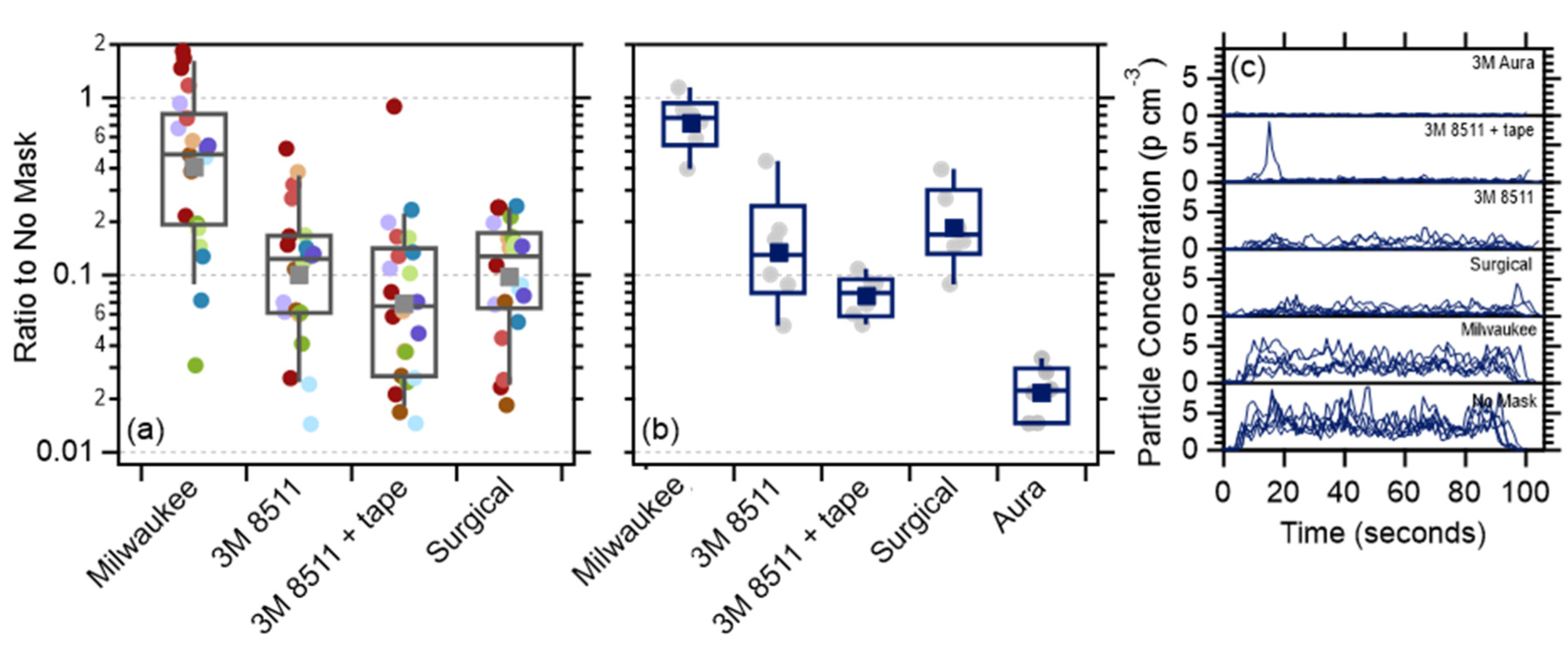
Observations of the reduction in respiratory particles emitted during speaking with wearing of various mask types. (a) Results for all participants, with colored points corresponding to different individuals. (b) Results for one individual repeating the task with wearing of different masks of given types. The box and whisker plots show the median (horizontal line), 25th/75th percentile (boxes), and 10th/90th percentiles (whiskers), along with the geometric mean (square). (c) The observed time-series of CPC-measured respiratory particle concentrations produced from speaking associated with the data in (b). Note that these have not been corrected for dilution.

The magnitude of the decrease in particle emissions with surgical mask wearing is consistent with our previous findings,^15, 16^ albeit with a somewhat higher overall efficiency. The *R*_mask_ of the 3M 8511 was similar to that observed by Asadi et al. (2020)^14^ for a different valved N95 respirator and in line with the range observed by NIOSH,^13^ while that for the Milwaukee was notably lower. Taping over the valve in the mask interior reduced the respiratory particle emissions by about a factor of two. The observed surgical mask efficiency was similar to the 3M 8511 mask and significantly better than the Milwaukee mask (*p* < 10^−4^). The trials by the participant who repeated the tasks multiple times additionally indicate that wearing of the non-valved 3M Aura N95 mask provided excellent reduction in exhaled particle concentrations, with *R*_mask_ = 0.022 (*Figure 2*b).

For a few participants, the particle concentrations with mask wearing exceeded that with no mask, which can occur when e.g., skin-mask rubbing releases non-respiratory particles.^16, 17^ Alternatively, this could reflect natural variability in the emission of respiratory particles by individuals; for the participant who repeated the tasks multiple times the ratio between the maximum and minimum observed particle emission rates equaled 1.6. The potential for non-respiratory particle contributions means that the actual reduction afforded by the masks could be greater than the observations suggest. However, we have no reason to think that the Milwaukee mask led to significantly greater production of such non-respiratory particles than the other masks as the fit and material was generally similar to that 3M 8511.

The individual *R*_mask_ for a given mask type varied widely between participants for all mask types but most notably for the Milwaukee mask. In general, the variability in both the absolute concentrations and the *R*_mask_ between participants (*Figure 2*a) greatly exceeded the difference in the replicates for an individual participant (*Figure 2*b), consistent with previous observations for surgical mask wearing.^15, 16^ The greater variability between individuals could indicate greater consistency in either how the masks were worn or in the speaking activity performed by one participant than between participants.

The Milwaukee and 3M 8511 respirators both have their valve similarly positioned in the center. As such, the very different performance of these two valved respirators likely results from a difference in the ease with which the valve opens during speaking. This indicates that highly variable performance of different valved respirators, if worn by the public, is expected and with some models providing almost no reduction in exhaled respiratory particles. Thus, although valved respirators can provide protection to the wearer against inhalation of endogeneous ambient particles, the use of valved masks when source control of respiratory particles is desired should be avoided, as is the case when the aim is reduction of respiratory disease transmission.

## Data Availability

All data produced in the present study are available upon reasonable request to the authors

## AUTHOR CONTRIBUTIONS

J.M.H. and C.D.C. conducted the experiments. C.D.C. conceived the research, analyzed the data, and wrote the paper with contributions from J.M.H.

## ACKNOWLEDGEMENTS

We thank all of the participants for their time and Prof. Bill Ristenpart (UC Davis) for use of the laminar flow hood.

## REFERENCES

1. U. S. Federal Drug Administration N95 Respirators, Surgical Masks, Face Masks, and Barrier Face Coverings. https://www.fda.gov/medical-devices/personal-protective-equipment-infection-control/n95-respirators-surgical-masks-face-masks-and-barrier-face-coverings (accessed 21 October 2021).

2. Oberg, T.; Brosseau, L. M., Surgical mask filter and fit performance. American Journal of Infection Control 2008, 36 (4), 276–282.

3. Lee, S.-A.; Grinshpun, S. A.; Reponen, T., Respiratory Performance Offered by N95 Respirators and Surgical Masks: Human Subject Evaluation with NaCl Aerosol Representing Bacterial and Viral Particle Size Range. The Annals of Occupational Hygiene 2008, 52 (3), 177–185.

4. Grinshpun, S. A.; Haruta, H.; Eninger, R. M.; Reponen, T.; McKay, R. T.; Lee, S.-A., Performance of an N95 Filtering Facepiece Particulate Respirator and a Surgical Mask During Human Breathing: Two Pathways for Particle Penetration. Journal of Occupational and Environmental Hygiene 2009, 6 (10), 593–603.

5. Shakya, K. M.; Noyes, A.; Kallin, R.; Peltier, R. E., Evaluating the efficacy of cloth facemasks in reducing particulate matter exposure. Journal of Exposure Science & Environmental Epidemiology 2017, 27 (3), 352–357.

6. Lee, S.-A.; Grinshpun, S. A.; Reponen, T., Respiratory performance offered by N95 respirators and surgical masks: Human subject evaluation with NaCl aerosol representing bacterial and viral particle size range. Annals of Occupational Hygiene 2008, 52 (3), 177–185.

7. Bellin, P.; Hinds, W. C., Aerosol penetration through respirator exhalation valves. American Industrial Hygiene Association Journal 1990, 51 (10), 555–560.

8. Lindsley, W. G.; Blachere, F. M.; Beezhold, D. H.; Law, B. F.; Derk, R. C.; Hettick, J. M.; Woodfork, K.; Goldsmith, W. T.; Harris, J. R.; Duling, M. G.; Boutin, B.; Nurkiewicz, T.; Boots, T.; Coyle, J.; Noti, J. D., A comparison of performance metrics for cloth masks as source control devices for simulated cough and exhalation aerosols. Aerosol Science and Technology 2021, 55 (10), 1125–1142.

9. Czypionka, T.; Greenhalgh, T.; Bassler, D.; Bryant, M. B., Masks and Face Coverings for the Lay Public. Annals of Internal Medicine 2021, 174 (4), 511–520.

10. Asadi, S.; Wexler, A. S.; Cappa, C. D.; Barreda, S.; Bouvier, N. M.; Ristenpart, W. D., Aerosol emission and superemission during human speech increase with voice loudness. Scientific Reports 2019, 9 (1), 2348.

11. Morawska, L.; Johnson, G. R.; Ristovski, Z. D.; Hargreaves, M.; Mengersen, K.; Corbett, S.; Chao, C. Y. H.; Li, Y.; Katoshevski, D., Size distribution and sites of origin of droplets expelled from the human respiratory tract during expiratory activities. Journal of Aerosol Science 2009, 40 (3), 256–269.

12. Staymates, M., Flow visualization of an N95 respirator with and without an exhalation valve using schlieren imaging and light scattering. Physics of Fluids 2020, 32 (11), 111703.

13. NIOSH Filtering facepiece respirators with an exhalation valve: measurements of filtration efficiency to evaluate their potential for source control; By Portnoff L, Schall J, Brannen J, Suhon N, Strickland K, Meyers J., U.S. Department of Health and Human Services, Centers for Disease Control and Prevention, National Institute for Occupational Safety and Health: DHHS (NIOSH) Publication No. 2021–107, 2020.

14. Asadi, S.; Cappa, C. D.; Barreda, S.; Wexler, A. S.; Bouvier, N. M.; Ristenpart, W. D., Efficacy of masks and face coverings in controlling outward aerosol particle emission from expiratory activities. Scientific Reports 2020, 10 (1), 15665.

15. Cappa, C. D.; Asadi, S.; Barreda, S.; Wexler, A. S.; Bouvier, N. M.; Ristenpart, W. D., Expiratory aerosol particle escape from surgical masks due to imperfect sealing. Scientific Reports 2021, 11, 12110.

16. Cappa, C. D.; Ristenpart, W. D.; Barreda, S.; Bouvier, N. M.; Levintal, E.; Wexler, A. S.; Roman, S. A., A highly efficient cloth facemask design. Aerosol Science and Technology 2021, 1–17.

17. Asadi, S.; Cappa, C. D.; Barreda, S.; Wexler, A. S.; Bouvier, N. M.; Ristenpart, W. D., Efficacy of masks and face coverings in controlling aerosol particle emission from expiratory activities. Scientific Reports 2020, 10, 15665.

